# Facemask wearing in COVID-19 pandemic: Correlates and prevalence; A survey after COVID-19 second wave in Uganda

**DOI:** 10.1101/2023.10.16.23297124

**Authors:** Nelson Onira Alema, Christopher Okot, Emmanuel Olal, Eric Nzirakaindi Ikoona, Freddy Wathum Drinkwater Oyat, Steven Baguma, Denish Omoya Ochula, Patrick Odong Olwedo, Johnson Nyeko Oloya, Francis Pebalo Pebolo, Pamela Okot Atim, Godfrey Smart Okot, Ritah Nantale, Judith Aloyo, David Lagoro Kitara

**Author notes:** **Corresponding Author**: David Lagoro Kitara is a Takemi fellow of Harvard University and Faculty at Gulu University, Faculty of Medicine, Department of Surgery, P.0. Box 166, Gulu City, Uganda.

## Abstract

**Background:** The WHO and the US. CDC documented that facemask-wearing in public situations is one of the most important prevention measures that can limit the acquisition and spread of COVID-19. Considering this, WHO and US. CDC developed guidelines for using facemasks in public settings. This study aimed to determine correlates and prevalence of facemask wearing during COVID-19 pandemic among adult population of Northern Uganda.

**Methods:** We conducted a cross-sectional study on five hundred and eighty-seven adult population of northern Uganda. A single stage stratified, and systematic sampling methods were used to select respondents from twenty-four Acholi subregion’s health facilities. Data was collected in a face-to-face questionnaire interview with an internal validity of Cronbach’s α=0.72. A local IRB approved the study, and Stata 18 was used for data analysis at multivariable Poisson regression with a p-value set at ≤0.05.

**Results:** The most substantial findings from this study were the high prevalence of face mask-wearing in public among respondents [88.7%,95%CI:86%-91%]. At a multivariable Poisson regression analysis, we found that obese respondents were 1.12 times more likely to wear facemasks than those who were not, [adjusted Interval Rates Ratios, aIRR=1.12,95%CI:1.04-1.19;p<0.01], and respondent who agreed to the lockdown measures were 1.23 times more likely to wear facemasks during COVID-19 pandemic than those who did not, [aIRR=1.23, 95%CI:1.07-1.41;p<0.01]. Other sociodemographic characteristics such as sex, age, occupation, level of education, religion, tribes, marital status, nationality, race, and comorbidities were not statistically significant at 95% Confidence Intervals.

**Conclusion:** The most significant findings from this study were the high prevalence of face mask-wearing among adult community members in northern Uganda. The correlates of facemask wearing in public were the obese and respondents who agreed with the presidential directives on the lockdown measures. Although this was within acceptable prevalence rates, the strict enforcement of face mask-wearing by security forces raised concerns among many community members and human rights advocates. We recommend more studies on communities’ perspectives on the challenges and benefits of facemask-wearing during the COVID-19 pandemic.

## Introduction

Coronavirus disease 2019 (COVID-19) is a worrying respiratory tract disease resulting from severe acute respiratory syndrome coronavirus-2 (SARS-CoV-2) infection.^1^ As the World Health Organization (WHO) recommended, several pandemic response activities were instituted across the globe to curb the spread of the coronavirus (COVID-19).^2^ Social distancing (SD)/Physical distancing (PD) and wearing facemasks are critical non-pharmaceutical public health interventions that have proven effective in reducing the spread of SARS-CoV-2.^2,3^ Facemask-wearing in public situations has been documented as one of the most important prevention measures that can limit the acquisition and spread of COVID-19 by the WHO and the United States Centers for Disease Control and Prevention (CDC).^4,5^ In light of this, the WHO, and US. CDC developed guidelines for using facemasks in public settings.^4,5^.

Published studies have shown that wearing facemasks to control the spread of infectious diseases has several advantages, including simple operation, strong sustainability, high health benefits, and good health economic benefits.^6,7,8^ In addition, other studies have shown that facemasks used by the general public are of potentially high value in limiting community transmission of infectious diseases.^9,10,11,12^ Also, facemask wearing has been documented to curb viral transmission from asymptomatic individuals to the population, thus limiting the epidemic’s growth rate or the reproductive number of the virus.^12^

In order to limit community’s spread of COVID-19, community-wide use of facemasks has been encouraged as a good and viable prevention and control measure.^13,14^ This is because facemasks also serve as visible signals of a widely prevalent respiratory pathogen, SARS-CoV-2, and a tool that reminds people of the importance of other infection control measures, such as social distancing.^15^

Therefore, facemasks are symbolic in that, beyond being tools, they can increase healthcare workers’ perceived sense of safety, well-being, and trust in their healthcare systems.^15^ A few African studies have assessed the population’s perceptions and compliance with COVID-19 lockdown measures and the use of facemasks.^16^ Some studies have reported adequate COVID-19-related compliance among health workers, but others have found significant gaps among the population.^16^

As previously noted, handwashing with soap, water, sanitizers, and facemasks, limit human-to-human transmission of the virus when droplets are shed into the environment or on inanimate surfaces.^2,3^

In a study to determine the adherence level, Bob OA *et al.* found that only 29% of participants adhered to all COVID-19 preventive measures of interest in the first phase of the SARS-CoV-2 outbreak in Uganda.^17^ However, adherence to some measures was very high.^17^ They reported that nearly all participants (96%) stated frequent handwashing with soap, but only 33% reported wearing a facemask in public.^17^

However, more sensitization regarding the importance of facemask wearing in containing the COVID-19 pandemic, subsidies, and free masks for those who may not be able to afford are required.

Additionally, it was found that a population could stabilize the COVID-19 outbreak and halt the viral transmission by enforcing preventive measures, such as wearing facemasks, hand hygiene, and physical distancing.^18^ This finding was reported when the State of Arizona in the USA enforced the wearing of face masks and other preventive measures; COVID-19 cases were reduced by 75% in a month.^18^

In Uganda, there was a community transmission of COVID-19 which resulted in a geometrical spread of the virus across most districts with resultant public health, economic, and socio-political implications.^19^ The demographic characteristics and silhouettes of Ugandan population has a wide base with a very thin apex at about 86 years, meaning the majority of Ugandan population is young and less than 30 years (Figure 1).

**Figure 1:**
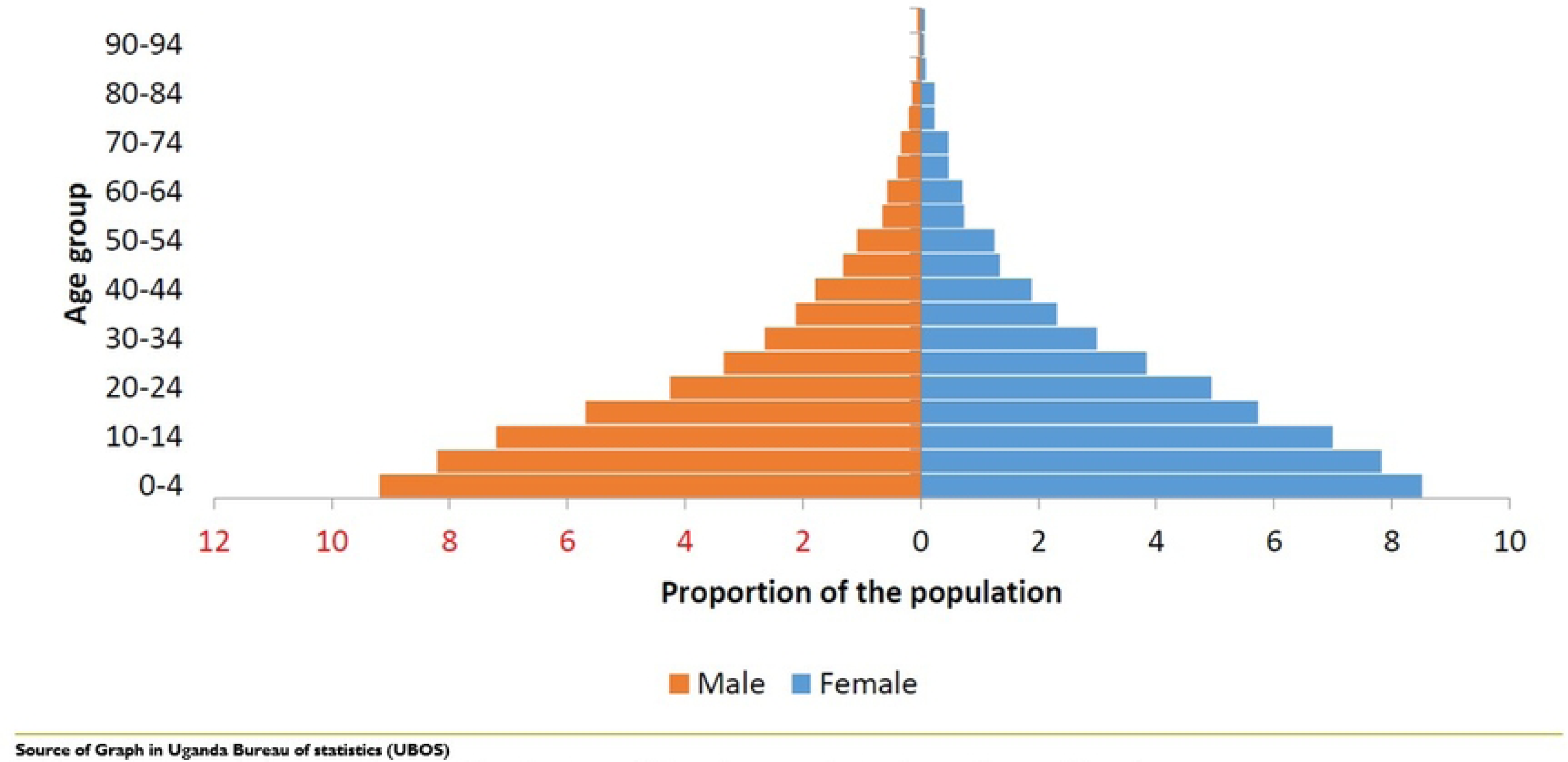
Age structure of Uganda’s population as of 2021, plotted by males versus females. In figure 1, a graph showing the age distribution of Ugandan population by males and females.

In northern Uganda, where this study was conducted (Figure 2), wearing facemasks before the COVID-19 pandemic was majorly a preserve of health workers in clinical and public health sectors and very rare in the general population. Indeed, face mask-wearing was not a common hygiene practice in the general population in the region. That being the case, it was necessary to determine the prevalence and correlates of facemask wearing among the general population of northern Uganda as a roll-out of key COVID-19 prevention measure with recommendations of the WHO, US. CDC and the Ugandan Ministry of Health.

**Figure 2:**
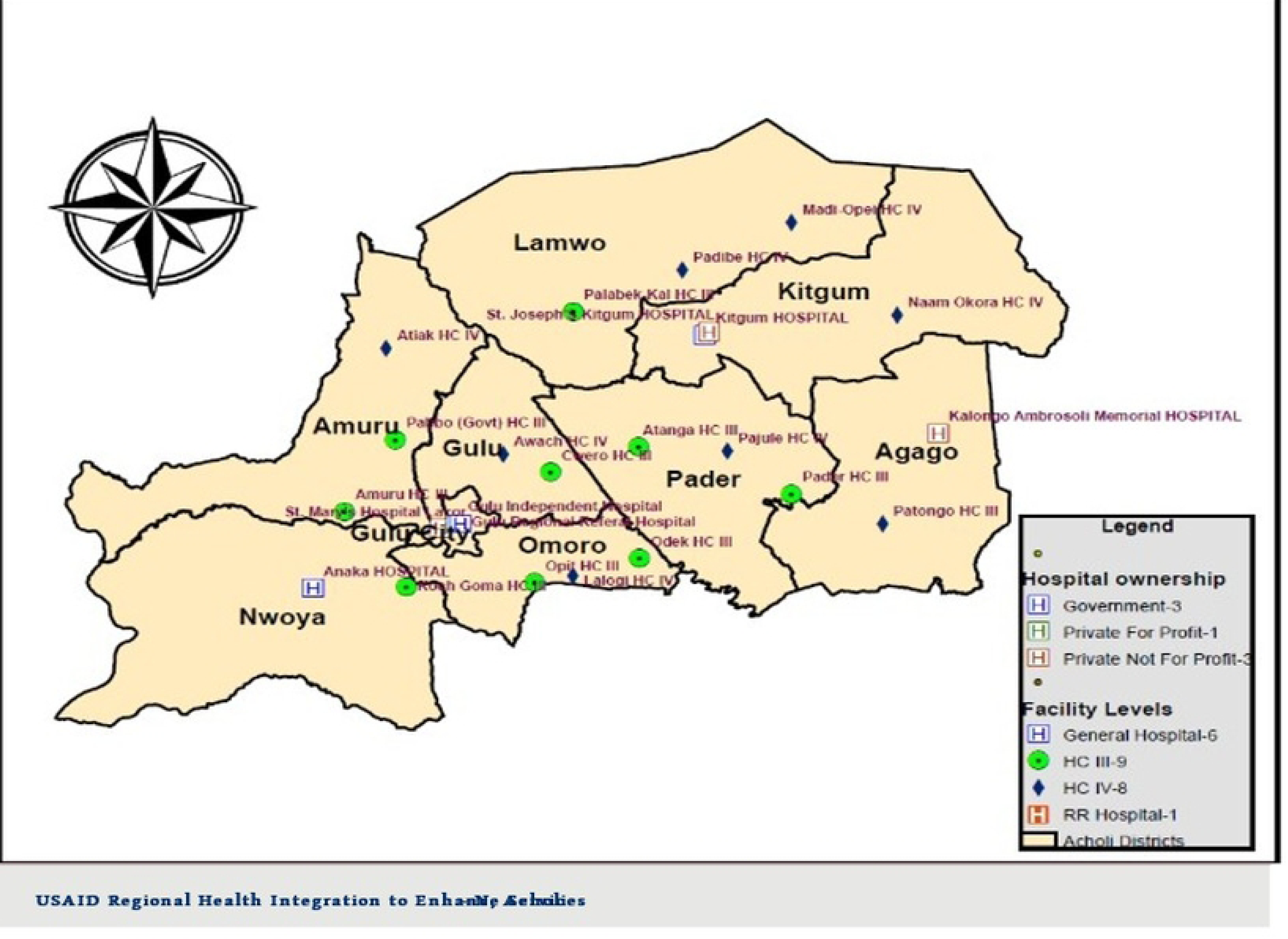
A map showing the study sites in the Acholi subregion. Figure 2 shows the study sites in northern Uganda.

This study aimed to determine the correlates and prevalence of facemask wearing during the COVID-19 pandemic among adult populations of northern Uganda.

## Methods

### Study design

We conducted a cross-sectional study in northern Uganda between October and November 2021.

### Study Sites

We conducted this study among adult community members of Northern Uganda in nine districts of the Acholi subregion (Gulu, Gulu City, Amuru, Nwoya, Omoro, Pader, Agago, Kitgum, and Lamwo districts).^20^ The nine districts are part of the Acholi subregion, which has just emerged from a 20-year-old war between the Government of Uganda and the rebel Lord’s Resistance Army (LRA), and the population is in the postwar period.^21^ In the nine districts, the total estimated population is two million, three hundred thousand people in a total land surface area of 28,500km^2^.^20,22,23^

During this study, Uganda had just eased the second lockdown measure to the COVID-19 second wave. The number of COVID-19 patients had significantly reduced in COVID-19 Treatment Centers (CTUs) in many health facilities in Northern Uganda.^24^ Health workers remained the frontline workforce (especially the nurses, doctors, and laboratory staff).^24,25^ In addition, district task forces set up by the Government of Uganda along the layers of administrative structures (national, districts, and communities) to support the management, prevention, and control of COVID-19 pandemic in communities met weekly to discuss new developments and plans of action.^24,25^ Further, the President of Uganda had announced new methods of work in public settings, whereby only 30% of staff in public and private organizations were allowed physically in offices.^24,25^ These COVID-19 control measures were intended to disrupt day-to-day contact between management, administration, and the community to interrupt the cycle of physical person-to-person contacts to break the transmission cycle of COVID-19.^24,25^

### Study participants and sampling techniques

We sampled five hundred and eighty-seven adult respondents recruited by a single stage stratified and systematic sampling techniques, and data was collected using a questionnaire. The questionnaire had two sections: Section A contained information on respondents’ sociodemographic characteristics and health background information (age, sex, occupation, tribe, religion, district, employment status, race, highest level of education, marital status, and habits such as smoking and drinking alcohol, and comorbidities such as obesity, Asthma, heart diseases, hypertension, Diabetes mellitus, and HIV and AIDS). In section B, we assessed the wearing of facemasks in public settings and perception on the lockdown measures instituted by the Government of Uganda.

We stratified the study population selection at regional level to the nine districts of the Acholi subregion. In the districts to the twenty-four health facilities where COVID-19 vaccination was carried to the general population without pay. Our study’s twenty-four selected health facilities (including Government and Non-governmental health facilities) included hospitals, Health Centre (HC) IVs, and Health Centre IIIs. The health facilities were Lacor Hospital, Gulu Regional Referral Hospital, independent hospital, Anaka Hospital, Kitgum Government Hospital, St. Joseph’s Hospital, Kitgum, and Dr. Ambrosoli Memorial Hospital, Kalongo (7 hospitals); Atiak HCIV, Lalogi HCIV; Pajule HCIV; Awach HCIV; Madi Opei HCIV, Padibe HCIV, and Namukora HCIV (7 Health Centre IVs); and Palabek HCIII, Amuru HCIII, Pabbo HCIII, Koch Goma HCIII, Opit HCIII, Pader HCIII, Patongo HCIII, Cwero HCIII, Odek HCIII, and Rackoko HCIII (10 HCIIIs).

In the health facilities’ outpatient departments, we conducted a systematic sampling technique on attendants and attendees of OPD from the OPD registers. We defined systematic sampling as a probability sampling method where researchers select population members at regular intervals.^26,27^ We chose this sampling technique because it allowed us to get the desired sample size in the shortest period, reducing the risk of our study team acquiring COVID-19.

Last but most importantly, a systematic sampling method helped to minimize biased samples and poor survey results in addition to eliminating clustered selection with a low probability of obtaining contaminated data^26,27^, which was the ideal situation the research team had to achieve.

### Sample size estimation

We calculated the sample size by using the Raosoft sample size calculator. The computation was on a 50% response distribution, 5% margin of error, and 95% Confidence Interval (CI). This online software foundation is a widely utilized descriptive sample size estimation formula.^28,29^ Based on the assumption of a total eligible population size of 50,000 (12.5% of the total adult above 18 years old in the Acholi subregion) in the nine districts of the Acholi subregion.

The minimum sample size based on the above assumptions and factoring in a 10% non-response rate was 437 participants.

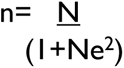

Where n= Sample size

N=the population size (50,000 people) e=margin of error at 5%

Substituting the formula

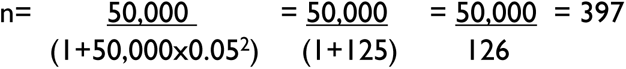

Add 10% for non-response, 39.7 +397 = 437

Based on the above assumptions, the minimum sample size after factoring a 10% non-response rate was 437. **Data collection**

We collected data using a pretested questionnaire designed by the research team. The pretest was conducted in the outpatient department of Gulu Regional Referral Hospital. The pretest results from the Gulu Hospital were not incorporated in the final data analysis. The questions achieved an internal validity of Cronbach’s α=0.72. After obtaining written informed consent from respondents, an interviewer-guided questionnaire was presented to respondents in a face-to-face questionnaire interview in the Outpatients’ department room, ensuring that infection, prevention, and control (IPCs) and standard operating procedures (SOPs) for COVID-19 were in place for both respondents and interviewers.^30^

First, OPDs were chosen as sites for this study because they had facilities for IPCs and SOPs. In addition, the OPD was the most convenient and preferred place to interview respondents as the population had just emerged from a severe second wave of COVID-19. During that period, the population was still in fear and apprehension due to the distress of contracting COVID-19 and were not willing to receive researchers in their offices, homes, or environment. Second, we adopted a face-to-face questionnaire interview as the best mode of data collection despite the risks of contracting COVID-19 because we wanted to reach out to as many participants as possible to answer our questionnaire. We could have used an online approach for data collection however, previous surveys conducted in northern Uganda showed very few online and internet users (23%)^31^ and mainly among persons who would not be eligible for this study because of age. Had we attempted to obtain data from online and internet users only, we would not have been able to obtain the sample size in time.

At each of the twenty-four selected health facilities, the study was in the Outpatients’ Departments (OPDs), where a consented adult person (≥18 years) was recruited for the study. The target population was attendees and attendants of the OPD services. A systematic sampling of every third attendant or attendee from the selected health facility’s OPD records was recruited from morning (9:00 am to 6:00 pm) every day from Monday to Saturday) until the sample size was achieved.

Each questionnaire interview with respondents lasted between 30-40 minutes in a convenient room in the OPD. As much as the questionnaire was in English, only a few respondents required translating some questions into the local language (5/587, 0.85%). We excluded respondents who could not speak (due to speech disability or inability to talk but not a language barrier) and were not residents of the Acholi subregion six months before the study. Interestingly, only two potential respondents declined to participate in the survey, constituting 2/589(0.34%) of the study population. Thus, the response rate for this study was 587/589(99.7%). Overall, the interviews went uneventfully for most respondents ensuring that standard IPC and SOP guidelines were adhered to.^30^

### Ethical approval

The study was approved by St. Mary’s Hospital, Lacor Institutional Review and Ethics Committee (LHIREC, No.0192/10/2021). In addition, this study followed institutional guidelines. We obtained written informed consent from each respondents aged (≥18 years). Furthermore, personal information of individual respondents was maintained confidential without including personal identifiers on the research documents. We kept all de-identified data under lock and key throughout the study period. Residual data was archived in the Department of Surgery, Faculty of Medicine of Gulu University.

### Data analysis

We analyzed this data using Stata 18^32^ and used Microsoft Excel 2019 to generate graphs. We conducted a descriptive analysis of respondents’ sociodemographic characteristics, presenting findings as proportions and percentages. We assessed the prevalence of facemask wearing among respondents and presented findings as frequencies and bar charts.

From the literature on COVID-19 facemask wearing during COVID-19 pandemic, we selected independent variables such as age, sex, occupation, level of education, employment status, race, nationality, tribes, religion, districts, addresses, comorbidity, smoking and drinking status, and marital status. The dependent variable was compliance with facemask wearing in public, defined as respondents reporting fidelity to wearing facemasks always in public and recorded as “yes” (1) or “no” (0).

We then applied a univariable modified Poisson regression to examine the relationship between each independent and dependent variable. Results were presented as crude Interval Rate Ratios (Crude IRR) and their respective P values at 95% Confidence Intervals (CI). We checked the collinearity between the independent and dependent variables using the variance inflation factor (VIF) method, and all VIF values were below two (2), indicating no evidence of collinearity.

Next, we fitted a multivariable Poisson regression model by entering all the selected independent variables to determine the factors associated with facemask wearing among the study population. The results were reported as adjusted Interval Rate Ratios (aIRR) with their respective P values and 95% Confidence Intervals. We considered a p-value ≤0.05 as statistically significant.

## Results

We conducted a study on adult population in the Acholi subregion and the questionnaire response was 587/589(99.7%) and only five respondents, 5/587(0.85%) required additional translation of the questionnaire from English to Acholi and two respondents declined to participate in the study 2/589(0.34%).

### Study sites

As seen in the map of the Acholi subregion in northern Uganda, the twenty-four health facilities are distributed according to the administrative structures in Uganda (Figure 2).

### Prevalence of facemask wearing in the study population

This study presents a unique finding where a community that does not usually wear facemasks faced the COVID-19 pandemic when there was a need to adjust behaviors and accept constraints to overcome the virus. Besides health workers in clinical and public health sectors, wearing facemasks was rare in the general population of northern Uganda until the COVID-19 pandemic struck. However, because of the guidance from the Ugandan Ministry of Health and Presidential directives on wearing facemasks in public, the population quickly accepted it and complied with the guidelines. The most substantial findings from this study were that the prevalence of facemask wearing among participants in public was high at [88.7%,95%CI:86%-91%] (Figure 3).

**Figure 3:**
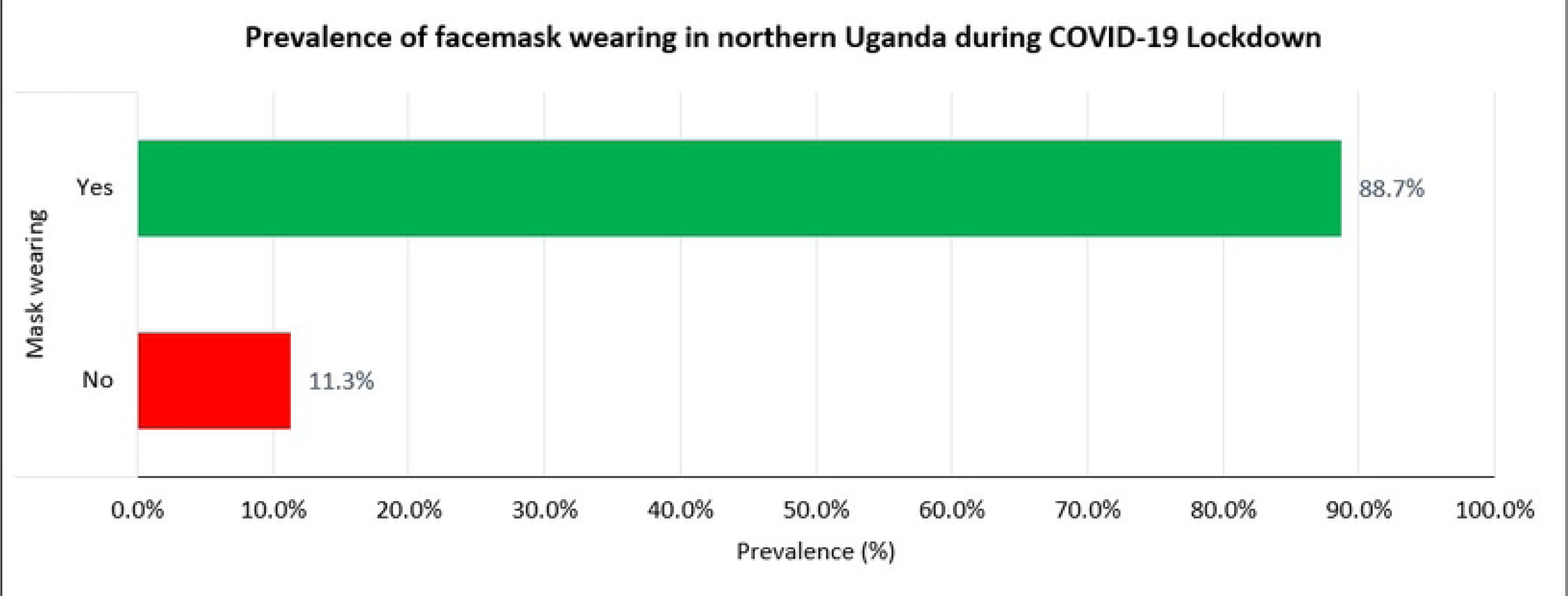
The prevalence of facemask wearing during COVID-19 lockdown in northern Uganda. In figure 1, 66(11.3%) reported not wearing facemasks, while 518(88.7%) reported always wearing facemasks as per national guidelines.

### Respondents’ characteristics

The study population was recruited from the twenty-four health facilities in the Acholi subregion of northern Uganda (Figure 2). A total of 587 respondents were enrolled in this study. The questionnaire response rate for this study was 587/589(99.7%). Most participants were males, constituting more than half of the population, 335(57.1%), and mainly in the age group of 25-34 years, 180(31.4%), who were either married or cohabiting, 341(58.9%). Also, most were Christians (94.9%), with Catholics being the predominant religion with over fifty percent, 312(53.2%). Regarding the tribes, the study area is predominantly Acholi ethnic groups, 425(72.9%), and were from the districts of Gulu/Omoro, 220(37.5%). Although information from the Uganda Demographic Health Surveys (UDHS)^22^ and Uganda Bureau of Statistics (UBOS)^23^ show that most population of the Acholi subregion had an adult literacy index of about 54%, most of our respondents had attained a tertiary level of education, 261(44.5%) (Table 1). They were employed in the informal sector 149(25.4%). Whereas there are several refugees, tourists, traders, and visitors in northern Uganda, most of our study respondents were Ugandans, 581(99%) who did not use alcohol, 401(69.0%); did not smoke cigarettes, 545(94.1%); had no diabetes, 571(97.3%); had no heart diseases, 571(97.3%); were not obese, 578(98.5%); had no hypertension, 559(95.2%); had no Asthma, 572(97.4%); were not HIV positive, 577(98.3%); and had no other chronic diseases, 542(92.3%) (Table 1).

**Table 1.**
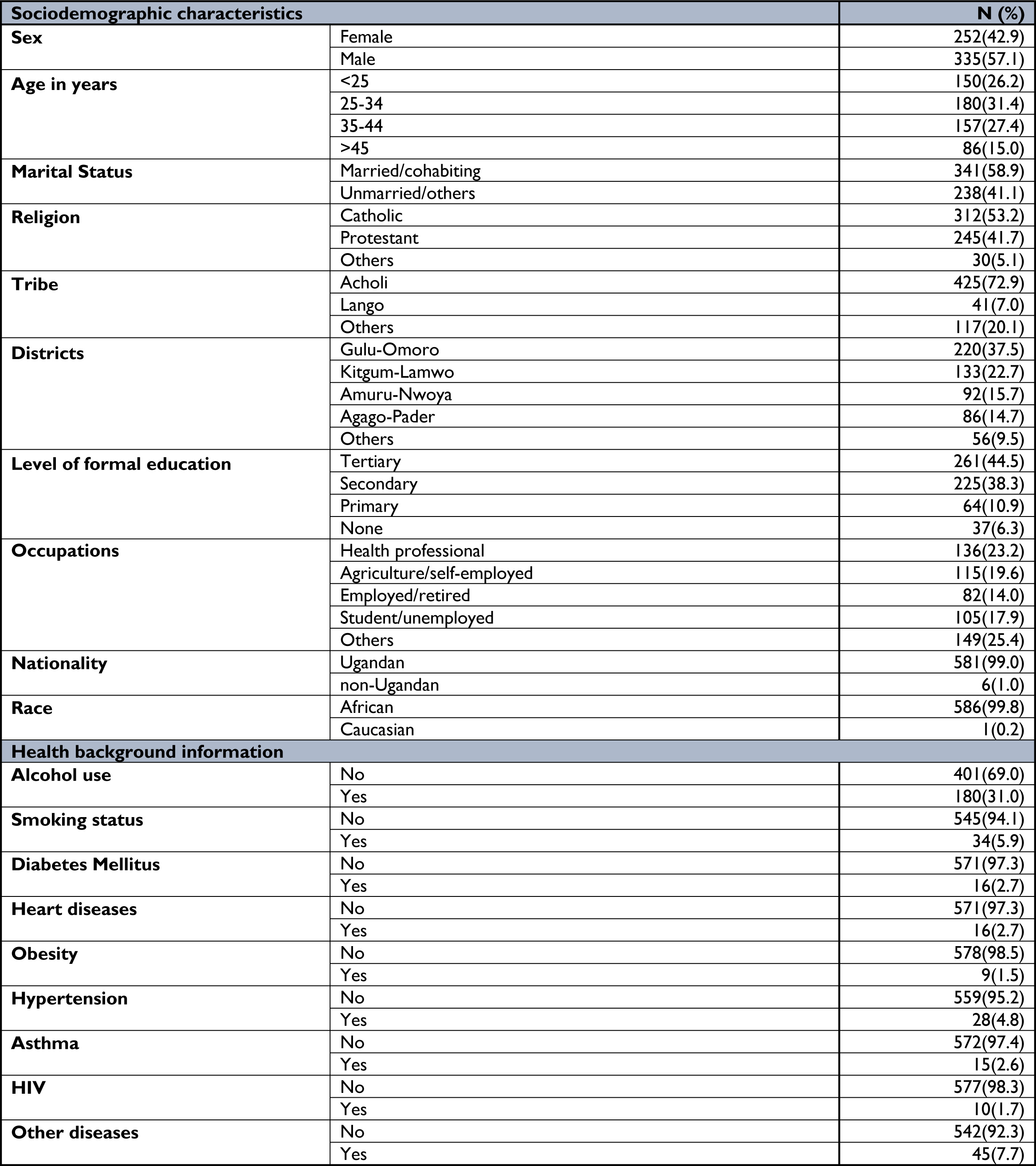
Sociodemographic and health background characteristics of respondents. In Table 1, most respondents were males, 335(57.1%); age-group of 25-34 years, 180(31.4%); married/cohabiting, 341(58.9%); Catholics, 312(53.2%); Acholi 425(72.9); from Gulu/Omoro districts, 220(37.5); with tertiary level of education, 261(44.5%); other occupations, 149(25.4%); Ugandans, 581(99%); did not use alcohol, 401(69.0%); did not smoke cigarettes, 545(94.1%); had no diabetes, 571(97.3%); had no heart diseases, 571(97.3%); were not obese, 578(98.5%); had no hypertension, 559(95.2%); had no Asthma, 572(97.4%); were not HIV positive, 577(98.3%); and no other diseases, 542(92.3%).

### Facemask wearing at bivariate analysis

To study the relationship between the dependent (wearing facemasks) and independent variables (sociodemographic and health background characteristics), we conducted bivariate analyses in which each independent variable was tested against the dependent variable. We found that the following variables were statistically significant at bivariate analysis, at 95% Confidence Intervals (CI) and p<0.05; the unmarried respondents had a [Crude Interval Rates Ratio, CIRR=1.06,95%CI:1.00-1.12;p=0.050]; obese respondents [CIRR=1.10,95%CI:1.10-1.16;p<0.010]; and respondents who agreed to the lockdown measures, [CIRR=1.22, 95%CI:1.07-1.39;p<0.010] (Table 2).

**Table 2:**
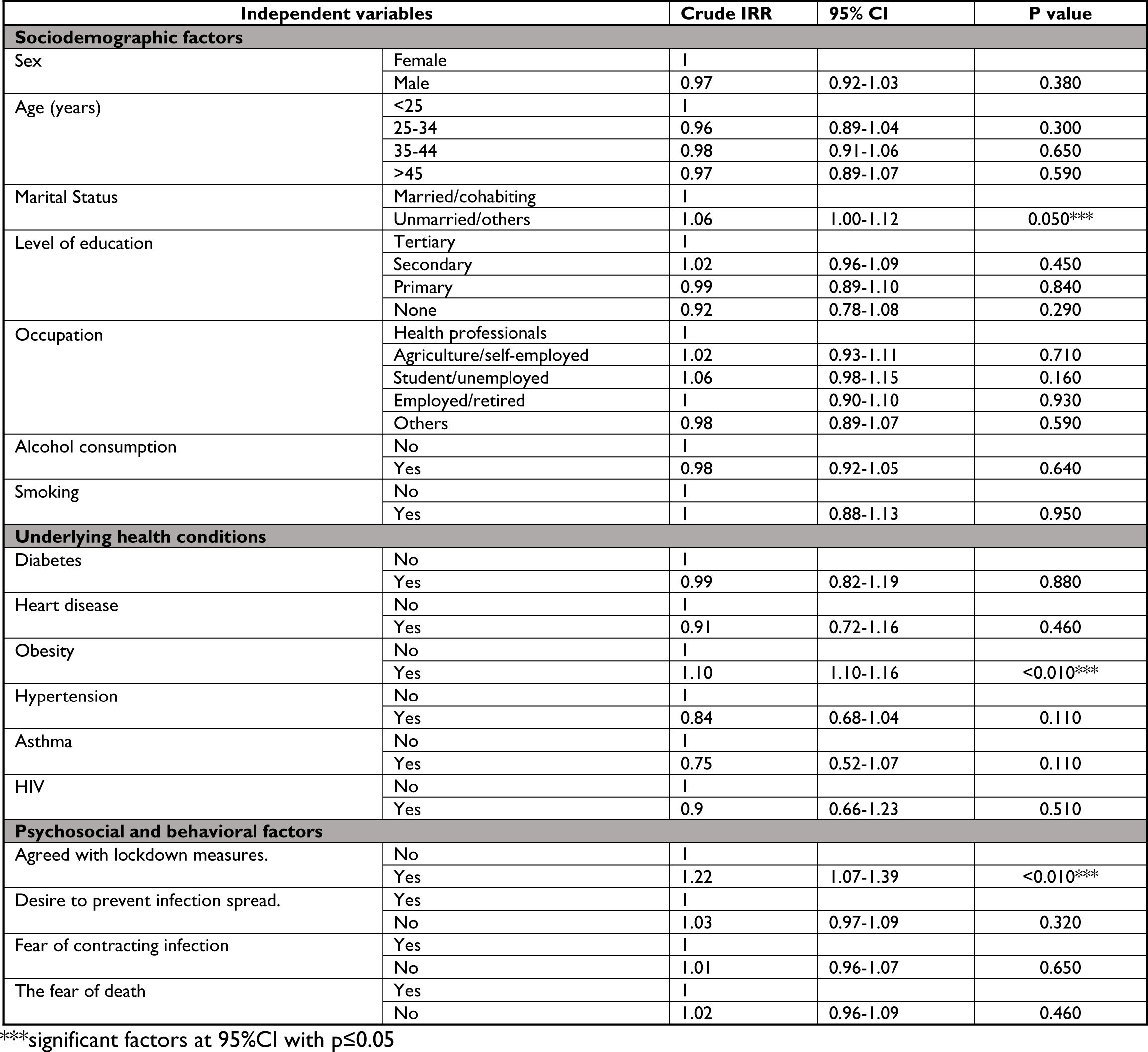
Factors associated with facemask wearing in Northern Uganda at bivariate analysis. Table 2 shows the factors that were significantly associated with facemask wearing at bivariate analysis; the unmarried with [Crude Interval Rates Ratio, CIRR=1.06, 95%CI:1.00-1.12;p=0.050]; Obese participants, [CIRR=1.10,95%CI:1.10-1.16;p<0.010]; and participants who agreed to the lockdown measures, [CIRR=1.22, 95%CI:1.07-1.39;p<0.010].

| Independent variables |  | Crude IRR | 95% CI | P value |
| --- | --- | --- | --- | --- |
| **Sociodemographic factors** |  |  |  |  |
| Sex | Female | 1 |  |  |
|  | Male | 0.97 | 0.92-1.03 | 0.380 |
| Age (years) | <25 | 1 |  |  |
|  | 25-34 | 0.96 | 0.89-1.04 | 0.300 |
|  | 35-44 | 0.98 | 0.91-1.06 | 0.650 |
|  | >45 | 0.97 | 0.89-1.07 | 0.590 |
| Marital Status | Married/cohabiting | 1 |  |  |
|  | Unmarried/others | 1.06 | 1.00-1.12 | 0.050*** |
| Level of education | Tertiary | 1 |  |  |
|  | Secondary | 1.02 | 0.96-1.09 | 0.450 |
|  | Primary | 0.99 | 0.89-1.10 | 0.840 |
|  | None | 0.92 | 0.78-1.08 | 0.290 |
| Occupation | Health professionals | 1 |  |  |
|  | Agriculture/self-employed | 1.02 | 0.93-1.11 | 0.710 |
|  | Student/unemployed | 1.06 | 0.98-1.15 | 0.160 |
|  | Employed/retired | 1 | 0.90-1.10 | 0.930 |
|  | Others | 0.98 | 0.89-1.07 | 0.590 |
| Alcohol consumption | No | 1 |  |  |
|  | Yes | 0.98 | 0.92-1.05 | 0.640 |
| Smoking | No | 1 |  |  |
|  | Yes | 1 | 0.88-1.13 | 0.950 |
| **Underlying health conditions** |  |  |  |  |
| Diabetes | No | 1 |  |  |
|  | Yes | 0.99 | 0.82-1.19 | 0.880 |
| Heart disease | No | 1 |  |  |
|  | Yes | 0.91 | 0.72-1.16 | 0.460 |
| Obesity | No | 1 |  |  |
|  | Yes | 1.10 | 1.10-1.16 | <0.010*** |
| Hypertension | No | 1 |  |  |
|  | Yes | 0.84 | 0.68-1.04 | 0.110 |
| Asthma | No | 1 |  |  |
|  | Yes | 0.75 | 0.52-1.07 | 0.110 |
| HIV | No | 1 |  |  |
|  | Yes | 0.9 | 0.66-1.23 | 0.510 |
| **Psychosocial and behavioral factors** |  |  |  |  |
| Agreed with lockdown measures. | No | 1 |  |  |
|  | Yes | 1.22 | 1.07-1.39 | <0.010*** |
| Desire to prevent infection spread. | Yes | 1 |  |  |
|  | No | 1.03 | 0.97-1.09 | 0.320 |
| Fear of contracting infection | Yes | 1 |  |  |
|  | No | 1.01 | 0.96-1.07 | 0.650 |
| The fear of death | Yes | 1 |  |  |
|  | No | 1.02 | 0.96-1.09 | 0.460 |

\*\*\*significant factors at 95%CI with $p \leq 0.05$

### Factors associated with facemask wearing in the study population

At multivariable poison regression analysis, we found that obese respondents were 1.12 times more likely to wear facemasks in public than those who were not, [Adjusted Interval Rates Ratios, aIRR=1.12,95%CI:1.04-1.19;p<0.01], and respondents who agreed to the lockdown measures during the COVID-19 pandemic were 1.23 times more likely to wear facemasks than those who did not, [aIRR=1.23, 95%CI:1.07-1.41;p<0.01]. Other sociodemographic and health background characteristics such as sex, age, occupation, level of education, religion, tribes, marital status, nationality, race, and comorbidities were not statistically significant at 95% Confidence Intervals (CI) (Table 3).

**Table 3:** Factors associated with facemask wearing in Northern Uganda during the COVID-19 pandemic at multivariable Poisson Regression Analysis. Table 3 shows the significant factors associated with facemask wearing during the COVID-19 pandemic in northern Uganda. These were; obese respondents, [adjusted Interval Rates Ratios, aIRR=1.12,95%CI:1.04-1.19;p<0.01], and participants who agreed with the lockdown measures, [aIRR=1.23, 95%CI:1.07-1.41;p<0.01].

| Independent Variables |  | Crude IRR | 95% CI | P value | Adjusted IRR | 95% CI | P value |
| --- | --- | --- | --- | --- | --- | --- | --- |
| **Sex** | Female | 1 |  |  | 1 |  |  |
|  | Male | 0.97 | 0.92-1.03 | 0.38 | 1.00 | 0.94-1.06 | 0.92 |
| **Age category (years)** | <25 | 1 |  |  | 1 |  |  |
|  | 25-34 | 0.96 | 0.89-1.04 | 0.30 | 0.98 | 0.90-1.07 | 0.71 |
|  | 35-44 | 0.98 | 0.91-1.06 | 0.65 | 1.03 | 0.95-1.11 | 0.54 |
|  | >45 | 0.97 | 0.89-1.07 | 0.59 | 1.02 | 0.92-1.12 | 0.73 |
| **Marital Status** | Married/cohabiting | 1 |  |  | 1 |  |  |
|  | Unmarried/others | 1.06 | 1.00-1.12 | 0.05 | 1.06 | 0.99-1.13 | 0.10 |
| **Obesity** | No | 1 |  |  | 1 |  |  |
|  | Yes | 1.1 | 1.10-1.16 | <0.01 | 1.12 | 1.04-1.19 | <0.01*** |
| **Agreed with lockdown measures** | No | 1 |  |  | 1 |  |  |
|  | Yes | 1.22 | 1.07-1.39 | <0.01 | 1.23 | 1.07-1.41 | <0.01*** |

\*Age and sex were taken as a priory in the multivariable analysis, \*\*\*significant factors at 95%CI with $p \leq 0.05$ .

## Discussion

Wearing facemasks in public venues where social distancing measures are problematic to maintain has been documented as one of the most important prevention measures that can limit the acquisition and spread of COVID-19 by the WHO and the US. CDC.^4,5^ Thus, the WHO and US. CDC developed guidelines for using facemasks in public environments during the COVID-19 pandemic.^4,5^ Epidemiological evidence has shown that community facemask wearing is effective in preventing the transmission of COVID-19.^33–36^ Our study from northern Uganda presented an optimal level of community facemask wearing during the COVID-19 pandemic at 88.7% (Figure 1). It showed that the level of community facemask wearing among adult community members in the Acholi subregion (Figure 2) during COVID-19 pandemic was acceptable (Figure 3) considering the postwar situation and higher prevalence of poverty in northern Uganda (using UBOS, 2020 data) compared to the rest of the county.^20,22,23^ Serial reports from the Uganda Bureau of Statistics (UBOS)^20,22,23^ showed that northern Uganda has consistently had worse poverty indicators (68%) than the rest of the country, and facemasks acquisition required money to purchase. Nevertheless, most Ugandan population needed help to afford facemasks during the pandemic which was distributed to a section of the population by the Government of Uganda (UBOS, 2020).^20^ Notably, the level of facemask wearing in our study was inconsistent with another study in Uganda by Mboowa *et al*.^37^ In their study, they reported a prevalence of facemask wearing of 70.3%, which was lower than ours at 88.7% (Figure 3). However, our level of facemask wearing was far higher than that of Amodan and colleagues at (33%)^38^ and lower than Ssebuufu *et al.* (99.3%).^39^ These differences could have been because Amodan and colleagues^38^ conducted their study at the earlier stages of the pandemic before initiatives such as mass distribution of facemasks, sensitization, mobilization, and engagement of communities^40^, and the strict enforcement of facemask use in public places^41,42,43^ was introduced in Uganda. Whereas the prevalence in Ssebuufu *et al.*^39^ was higher than ours, it did not use face-to-face interviews or guilt-free questions when assessing participants’ self-reported practices as we did.^44^

Therefore, findings on facemask wearing in Uganda may reflect the type of participants recruited and the period of the pandemic when the study was conducted. In addition, their participants were Ugandan community members who were recruited by online snowballing sampling techniques^39^, thus presenting potential risks of selection biases and non-probability sampling of respondents. Thus, the prevalence of face mask-wearing could vary according to the study respondents, knowledge levels, attitudes, practices, exposures, and risk perceptions.

This acceptable level of community facemask wearing among adult community members in northern Uganda emphasizes the need for initiatives to scale up the practice now and in the future to prevent and control any emerging infectious respiratory diseases (Table 3).

From a global health perspective, our findings show that the prevalence of face mask-wearing in public in northern Uganda was comparable to many communities in Asian countries where the practice is part of their cultures. For example, our study found that wearing facemasks during the COVID-19 pandemic in northern Uganda was much higher than in Hong Kong^45^, Malaysia^46^, and Ghana^47^ at 69.2%, 51.2%, and 32.3%, respectively. Ours was almost similar to studies in Uganda^48,49^ and Pakistan^50^ where they found that 99.3%, 93%, and 93.9% of respondents, respectively, reported wearing facemasks during the COVID-19 pandemic when in public.^48,49,50^

Many factors might explain these findings. First, the belief and practice that facemasks could protect them against COVID-19. Second, the security forces strict enforcement of facemask-wearing in Uganda gave no room for defaulting.^41,42,43^ Third, the government of Uganda supplied facemasks to community members across the country, so access to facemasks was not a problem. Fourth, because the nature and quality of the facemasks did not matter, some community members used clothes or handkerchiefs to cover their faces in public, and these were accepted as facemasks.

Also, our study assessed compliance and satisfaction with wearing facemasks as an appropriate COVID-19 prevention measure after Uganda’s second and most severe wave of COVID-19 outbreaks. 88.7% of our study population reported wearing facemasks in public settings during the pandemic (Figure 3). In addition, the level of satisfaction with facemask use was higher at (90%).

It was previously estimated that proper wearing of facemasks with a coverage of 80% would halt the transmission of the virus.^51^ This was confirmed when the State of Arizona in the USA enforced the wearing of facemasks and other preventive measures; COVID-19 cases were reduced by 75% in a month.^18^

However, like other countries in the African continent, facemask wearing is not commonly practiced and it was only introduced to the African communities in response to the COVID-19 pandemic.^52^ In addition, the low usage of facemasks in some communities in the African continent could have resulted from initial information needing to be more consistent about the importance of facemask wearing for the prevention of COVID-19 transmission by the general population.^52^ Additionally, there was information circulating in many communities that the threat posed by COVID-19 to Ugandan and African populations was likely mild considering the warm tropical environment, less crowded environment, and a predominantly young population structure.^53^ In contrast to our findings in northern Uganda, many communities in the African continent did not wear facemasks because they were uncomfortable or because they did not think they were necessary.^54^

Further, our study found that the correlates of facemasks wearing were respondents with comorbidity (obesity) and those who agreed with the lockdown measures instituted by the government of Uganda to curb the exponential spread of COVID-19 (Table 3). This finding is consistent with many studies that show that a population with high-risk perceptions were more likely to adhere to facemask wearing as a prevention measure for the control of infectious diseases such as COVID-19.^24,25,34,37,39^ In addition, the comorbid respondents were a special target group by the Ugandan Ministry of Health campaigns during the pandemic with specific emphasis on preventing the high-risk population from acquiring the virus, developing severe disease, hospitalization, and death.^24,25,34,37,39^ Furthermore, the Ugandan Ministry of Health prioritized the high-at-risk community members with supplies of facemasks, vaccines, and awareness campaigns on the dangers posed by COVID-19 to them.^24,25,34,37,39^

The high-risk perceptions among obese respondents in northern Uganda were rooted in the high COVID-19 mortality observed among obese patients seen in one of the health facilities in Uganda.^55^

A modeling study showed that comorbidity could impact the share of mild cases that develop severe symptoms.^56^ In Asia and Europe, hypertension, obesity, diabetes, and coronary heart diseases have been drivers of adverse health outcomes from COVID-19.^56,57^ The combined prevalence of diabetes, hypertension, and obesity was higher in regions that were used to derive the recovery rates when baseline simulations had already been accounted for compared to Ghana, Kenya, and Senegal where other comorbidities afflict them. We, the authors propose, further studies in order to reach a definite conclusion on the trending issue of comorbidity. For example, Ghana, Kenya, and Senegal have persistent and high rates of anemia and Tuberculosis (TB)^58^. However, there were no studies on the magnitude of the impact of anemia, TB, HIV, and AIDs on the recovery of patients who contracted COVID-19. Therefore, there is more to be considered as additional information about adverse consequences of comorbidity in different settings was being considered.

Because of many uncertainties about COVID-19, participants with comorbidities needed to consider protecting themselves by consistently wearing facemasks.

Finally, our findings that facemask wearing was more likely among respondents who agreed with the lockdown measures was consistent with many studies in Uganda.^59,60,61,62^ The lockdown measures for COVID-19 prevention and control were implemented in Uganda during the first and second waves of the pandemic.^59,60,61,62^ In both waves, lockdown measures were instituted through presidential directives strictly enforced by Uganda’s security forces.^42–43,59–62^ So, facemask wearing became mandatory when an individual was in public settings, and that could explain the high coverage in our study findings.^42–43^

In addition, northern Uganda is in a postwar era where the experience of war and the use of security forces to enforce government directives were fresh in the minds of the population.^21^ It is likely that some community members wore facemasks not because of conviction or belief that it was helpful but rather the fear that one would be apprehended or punished by the security forces.^24^

Inconsistent with our study findings, another in Somalia found that only half of their respondents reported wearing facemasks during the COVID-19 pandemic.^63^ This was far below the 80% threshold recommended by modeling studies in England and whales^64^, and therefore the need for facemask wearing to be improved in Somalia.^63^

Of particular note, reusable cloth facemasks were reported only in about one-quarter of the participants.^63^ Cloth facemasks, however, offer some advantages, such as lower costs and being far less dangerous for the environment when compared to surgical masks, and have been proposed for COVID-19 control, especially in resource-poor settings.^63,65^ Aside from the reported difficulties of acquiring masks (such as lack of finances and not knowing where to obtain masks), as also observed in our study, other significant limitations to wearing facemasks included the associated discomfort and the widespread thought that masks were unnecessary in times of COVID-19.

Indeed, face mask-wearing is a relatively strange practice among the African population, and they should be educated about its importance during this COVID-19 crisis. This finding contrasts with Asian countries, where masks are culturally accepted as a common hygiene practice.^66^

### Strengths and limitations of this study

This study had many strengths. First, it was conducted on a large sample size among community members in northern Uganda, so most of the information obtained can be generalizable. Second, we used a systematic sampling method which is a probability sampling method, and thus, the information obtained could represent findings in the specific location of similar settings. Third, the information obtained can help inform policy on facemask wearing in communities where masks are culturally not part of their hygiene practice.

However, this study had some limitations. First, the nature of the study design, a cross-sectional study with inherent limitations of not measuring variables over time, thus risk not capturing the dynamism of changing times and perceptions of respondents. Second, we captured the views and opinions of adult respondents ≥18 years, yet most of the population in northern Uganda were below 18 years (Figure 1). This finding presents a challenge of representation bias among younger age groups and may become problematic when designing strategies for preventing and controlling such diseases in future outbreaks. Finally, our findings that most respondents in our study had attained a tertiary level of education pose a challenge of representation. Information obtained from studies in northern Uganda shows that most of the population does not have tertiary education.^22^ This finding may present a selection bias in our study population.

### Generalizability of results

These findings are generalizable to rural communities in sub-Saharan Africa with similar contexts.

## Conclusions

The most significant findings from this study were the high prevalence of face mask-wearing among adult community members in northern Uganda. The correlates of wearing facemasks were obese respondents and those who agreed with the presidential directives on lockdown measures. Although this is within acceptable prevalence rates, the strict enforcement of the practice by security forces has raised concerns among many community members and human rights advocates. We recommend more studies on communities’ perspective on the challenges and benefits of facemask-wearing after the COVID-19 pandemic.

## Abbreviations

AIDS: Acquired immune deficiency syndrome
HIV: Human immune virus
HC: Health Centre
CI: Confidence intervals
COVID-19: Coronavirus disease-19
LHIREC: Lacor Hospital Institutional Review and Ethics Committee
IPC: Infection, prevention, and control
SOPs: Standard Operating Procedures
WHO: World Health Organization
US. CDC: United States of America Centers for Disease Control and Prevention
UBOS: Uganda Bureau of Statistics
UDHS: Uganda Demographic Health Surveys
WHO: World Health organization.

## Declarations

### Ethics approval and consent to participate

This study was approved by St. Mary’s Hospital Institutional Review and Ethics Committee (LHIREC, No.0192/10/2021). In addition, the study followed all relevant institutional guidelines and regulations. We obtained written informed consent from each respondent and their legal representatives in this study.

### Consent to publish

We obtained informed consent from participants to publish this information.

### Availability of data and material

All datasets supporting this article’s conclusion are within this paper and are accessible by a reasonable request to the corresponding author.

### Authors’ contributions

DLK, JA, and FWDO designed this study—JA, FWDO, POA, SB, CO, GSO, FPP, and DLK supervised data management. ENI, JA, FWDO, EO, RN, and DLK analyzed and interpreted the data. FPP, NOA, POA, CO, DOO, GSO, EO, ENI, FWDO, POO, JNO, BS, RN, JA, and DLK wrote and revised the manuscript. All Authors approved the manuscript.

### Authors’ Information

Dr. Nelson Onira Alema (NOA) is a Lecturer at Gulu University, Faculty of Medicine, Department of Anatomy, Gulu City, Uganda; Dr. Christopher Okot (CO) is a Medical Officer Special-Grade, Department of Surgery at Gulu Regional Referral Hospital, Gulu City, Uganda; Dr. Emmanuel Olal (EO) is a Public Health Specialist and a Physician at Yotkom Medical Centre in Kitgum District, Kitgum, Uganda; Dr. Eric Nzirakaindi Ikoona (ENI) is a Technical Director at ICAP at the University of Columbia, Sierra Leone; Dr. Freddy Wathum Drinkwater Oyat (FWDO) is a Senior Physician, a Public Health Specialist, and a member of Uganda Medical Association, UMA-Acholi branch, Gulu City, Uganda; Dr. Steven Baguma (SB) is a Medical Officer in the Department of Obstetrics and Gynecology at Gulu Regional Referral Hospital, Gulu City, Uganda; Dr. Denish Omoya Ochula (DOO) is a District Health Officer at Lamwo district local government, Lamwo, Uganda; Dr. Patrick Olwedo Odong (POO) is a Hospital Director at Yumbe Regional Referral Hospital, Yumbe district, Uganda; Dr. Johnson Nyeko Oloya (JNO) is a Medical Officer at Moroto Regional Referral Hospital, Moroto district, Uganda; Dr. Francis Pebalo Pebolo (FPP) is a Lecturer at Gulu University, Faculty of Medicine, Department of Reproductive Health, Gulu City, Uganda; Dr. Pamela Okot Atim (POA) is a Senior Obstetrician and Gynecologist and a Medical Superintendent at St. Joseph’s Hospital, Kitgum, Uganda; Dr. Geoffrey Smart Okot (GSO) is a Senior Surgeon and a Medical Superintendent at Dr. Ambrosoli Memorial Hospital, Kalongo, Agago district, Uganda; Dr. Ritah Nantale (RN) is a Research Manager for the Centre of Excellence for Maternal, Reproductive and Child Health at Busitema University, Mbale City, Uganda; Dr. Judith Aloyo (JA) is a Deputy Chief of Party at Rhites-N, Acholi, Gulu City, Uganda; Prof. David Lagoro Kitara (DLK) is a Takemi fellow of Harvard University and a Professor at Gulu University, Faculty of Medicine, Department of Surgery, Gulu City, Uganda.

## Acknowledgment

We thank the assistance from health facilities for the datasets obtained and Dr. David Mukunya for the comprehensive data analysis of this study.

